# Cancer risk and survival outcomes in germline *POT1* variant carriers

**DOI:** 10.64898/2026.11.10.26367070

**Authors:** Anna C. Raper, Baergen I. Schultz, Jung Kim, Bradley S. Wubbenhorst, Annika Martin, Mersedeh Rohanizadegan, Tiffiney R. Hartman, Stacey Cohen, Teresa Chai, Aniket Dagar, Jeremy Haley, Jiang Li, David Carey, Regeneron Genetics Center, Penn Medicine Biobank, Dirk Hockemeyer, Douglas R. Stewart, Katherine L. Nathanson

## Abstract

**Background:** The prevalence of germline *POT1* variants, spectrum of associated tumors, cancer risk and survival outcomes are incompletely described, and clinical management guidelines vary. We evaluated *POT1* variant carriers in three biobanks and a clinical cohort to address these limitations.

**Methods:** Prevalence, cancer spectrum and risk associated with germline *POT1* pathogenic/likely pathogenic and additional functionally assessed variants were evaluated in Penn Medicine Biobank (PMBB; *n*=51,172), Geisinger MyCode (MyCode; *n*=220,582), UK Biobank (UKBB; *n*=469,558) and a Penn Medicine clinical cohort.

**Results:** Estimated *POT1* variant prevalence ranged from 1:1,279 to 1:2,902. Compared to non-carriers, cancer diagnoses were more frequent among *POT1* variant carriers in PMBB (OR 3.41, 95%CI 1.81-6.43, *p*=1.88x10^-4^), MyCode (OR 2.23, 95%CI 1.31-3.79, p=0.003), and UKBB (OR 1.40, 95%CI 1.07-1.84, *p*=0.015). Cancer incidence increased beginning in the mid-30s, without evidence of increased mortality. Along with previously described cancer types, lung cancer was associated with *POT1* variant carriers in two biobank cohorts.

**Conclusion:** *POT1* is a cancer susceptibility gene associated with a moderate (1.5-3.5-fold) increased cancer risk. Variants are relatively frequent and predispose to a broad tumor spectrum and increased incidence starting in the mid-30s, without increasing mortality. These findings expand understanding of *POT1*-associated disease and provide data to inform clinical management.

## INTRODUCTION

Protection of Telomeres 1 (POT1) protein is a highly conserved subunit of the shelterin telomere binding complex, which binds and protects human telomere ends and regulates telomere length.^1–3^ Loss of POT1 function causes unprotected telomere ends, leading to aberrant telomere lengthening and chromosomal abnormalities, which promotes tumorigenesis.^4–7^ Germline *POT1* pathogenic variants (PVs) associated with cancer predisposition were first identified in high-risk melanoma families, and melanoma remains the most commonly reported malignancy.^8,9^ However, *POT1* PVs have since been described in families and cohorts with glioma, angiosarcoma, colorectal, prostate, lung, and non-medullary thyroid cancers, as well as lymphoid and myeloid malignancies, particularly chronic lymphocytic leukemia (CLL).^10–22^ These reports have largely relied on small, clinic-based or disease-specific cohorts, limiting phenotypic breadth and precluding reliable cancer risk estimation.

Cancer risk associated with *POT1* PVs in the general population has not been characterized. A clinical testing cohort of 227 individuals with *POT1* PVs were significantly more likely to develop multiple tumors, particularly melanoma and sarcoma.^23^ A pan-cancer cohort of 23 patients reported increased risks of papillary thyroid cancer and CLL, as well as younger age at cancer diagnosis.^24^ These two retrospective studies using cohorts with prior genetic testing had limited phenotyping, and may provide inflated risk estimates, as the calculated odds ratios for cancer risk from cohorts with genetic testing for clinical reasons can be substantially higher than population-based cohorts.^25^ In addition to reporting on a clinical cohort, one of the aforementioned studies evaluated hematologic malignancy risk in the UKBB, which found higher lymphoid malignancy rates in *POT1* variant carriers^22^. Another study evaluated prevalence of pathogenic variants in familial melanoma genes, including *POT1*, only evaluating known cancer associations, and replicating them in cutaneous melanoma, hematologic malignant neoplasms, and thyroid cancer.^26^

The limited data on cancer risk, and lack of data on time to cancer onset and mortality in *POT1* PV carriers, have contributed to markedly discrepant clinical guidelines. One guideline recommends intensive surveillance resembling that for Li-Fraumeni syndrome, including melanoma surveillance, CBC, annual whole-body and brain imaging starting at age 18.^27^ In contrast, a UK expert group recommended surveillance only for melanoma, citing insufficient evidence for broader surveillance.^28^ Given increasing identification of *POT1* variants through multigene panel testing, population-based data are urgently needed to inform evidence-based management. Additionally, knowledge of pathogenicity of *POT1* variants remains limited, although recent functional studies have provided data on variants that are not confirmed pathogenic or likely pathogenic, but result in reduced POT1 function. Herein, we describe the prevalence of *POT1* P/LPVs, spectrum of malignant neoplasms, cancer risk and survival outcomes in individuals with germline *POT1* P/LPVs from three biobanks and a clinical cohort, coupled with an expanded understanding of pathogenic variants based on functional studies.

## METHODS

### Cohorts

Participants carrying *POT1* variants were identified from three biobank cohorts and a genetics clinic. Demographic and clinical variables are described in **Supplementary Methods.**

### Penn Medicine Biobank (PMBB)

PMBB is an electronic health record (EHR)-linked biobank of genomic and other biomarker data.^29^ PMBB includes 51,172 individuals with exome sequence data performed in collaboration with the Regeneron Genetics Center. This study was carried out under IRB protocol 832122.

### Geisinger MyCode Community Health Initiative

The DiscovEHR study consists of a subset of individuals who consented to participate in the Geisinger MyCode Community Health Initiative.^30^ Exome sequencing on 220,582 participants with EHR data was performed in collaboration with the Regeneron Genetics Center.

### United Kingdom Biobank (UKBB) Population-based Cohort

UKBB participants with *POT1* variants were ascertained from a population-based cohort study of 469,558 participants with whole exome sequencing (UKBB projects 69929, 54389) with linkage to electronic health records and the UK National Cancer Registration and Analysis Service (NCRAS).^31^ The UKBB is approved by the UK Biobank Research Ethics Committee.

### Penn Medicine Clinical Genetics Patients (Clinical Cohort)

Twenty-one patient-participants from 18 families with *POT1* variants identified clinically were evaluated between May 2017 and September 2025. Patients had genetic testing identifying a *POT1* variant based on either a personal or family history of cancer or a known familial *POT1* variant. Chart review was carried out under IRB protocol 854086. The clinical patients did not overlap with the PMBB identified patients.

### Variant annotation and classification

Two types of *POT1* variants were considered in this analysis: 1) pathogenic/likely pathogenic variants (P/LPV) including the common variant c.1851_1852del, p.Asp617fs; and 2) missense variants which substantially impair POT1 function, which are referred to as “reduced function missense” (RFM) variants (**Supplementary Table 1**). Variants identified in the biobank cohorts were classified as P/LPV using the Variant Effect Predictor (VEP) and AutoGVP,^32^ based on calls in ClinVar.^33^ Although p.Asp617fs is not expected to cause nonsense mediated decay, multiple lines of evidence support pathogenicity including downstream variants co-segregating with disease, functional evidence (**Supplementary Figure 1**); it is called likely pathogenic or pathogenic by three groups in ClinVar (SCV000655168.10). RFM variants were determined by allele depletion studies in deep mutation scanning in locally haploid human stem cells as previously described.^4,34–37^ One clinical cohort participant with a *POT1* variant of uncertain significance (c.817C>T, p.R273W) was included, as the variant was determined to be an RFM. This patient’s personal and family history also were consistent with *POT1* tumor predisposition (Family 1, **Supplementary Figure 2**). Analyses were conducted for P/LPVs (including p.Asp617fs variant carriers), and P/LPV + RFM.

### Phenotypic data

For *POT1* variant carriers identified in the biobank cohorts (PMBB, MyCode and UKBB), cancer diagnoses were extracted using ICD-9/10 codes and cancer registry data specific to each biobank, requiring at least two ICD code instances. Details are in **Supplementary Methods**. Cancer, family history and other medical history phenotypes were characterized in individuals with *POT1* P/LPVs or RFMs from the PMBB and the clinical cohort via EHR review at Penn Medicine. Patient identifiers used in supplemental materials were not known to anyone outside of the research group and are not identifying. Individuals with known germline P/LPVs in additional cancer predisposition genes were excluded from analysis. Permission from four clinical cohort families to publish their modified family history was obtained (**Supplementary Figure 2**).

### Statistical analyses

Continuous variables are presented as mean (SD); categorical variables as counts and percentages. Group comparisons used t tests, Mann–Whitney U tests, or Fisher exact tests, as appropriate, details in **Supplementary Methods**. Logistic regression models estimated cancer risk odds ratios (ORs) with adjustment for year of birth, sex, smoking history, and body mass index. Time-to-event analyses used Cox proportional hazards models adjusted for year of birth, sex, smoking history, ancestry, and body mass index. Odds ratios and hazard ratios are reported with 95% confidence intervals (CIs) and exact *P* values. Statistical significance was defined as 2-sided *P* < .05. Analyses were conducted using R version 4.1.0 and SAS Enterprise Guide version 8.3.

## RESULTS

### Prevalence and distribution of germline P/LP variants in POT1

We considered two groups of variants in *POT1*: 1) P/LPVs including c.1851_1852del (p.Asp617fs); and 2) P/LPVs and reduced function missense variants as determined in previous allele depletion studies^4,34–37^ (RFM; **Supplementary Table 1**). Across biobanks, the prevalence of *POT1* germline P/LPVs ranged from 1:1,279 to 1:2,902 (**Figure 1a**); with the inclusion of RFM, the rate ranged from 1:1,163 to 1:2,398. Variants were distributed throughout the *POT1* gene, predominantly within known functional domains (**Figure 1b**). The p.Asp617fs variant was the most frequent, accounting for 6.8% of PMBB, 18.5% of MyCode, and 25.4% of UKBB *POT1* variants.

**Figure 1:**
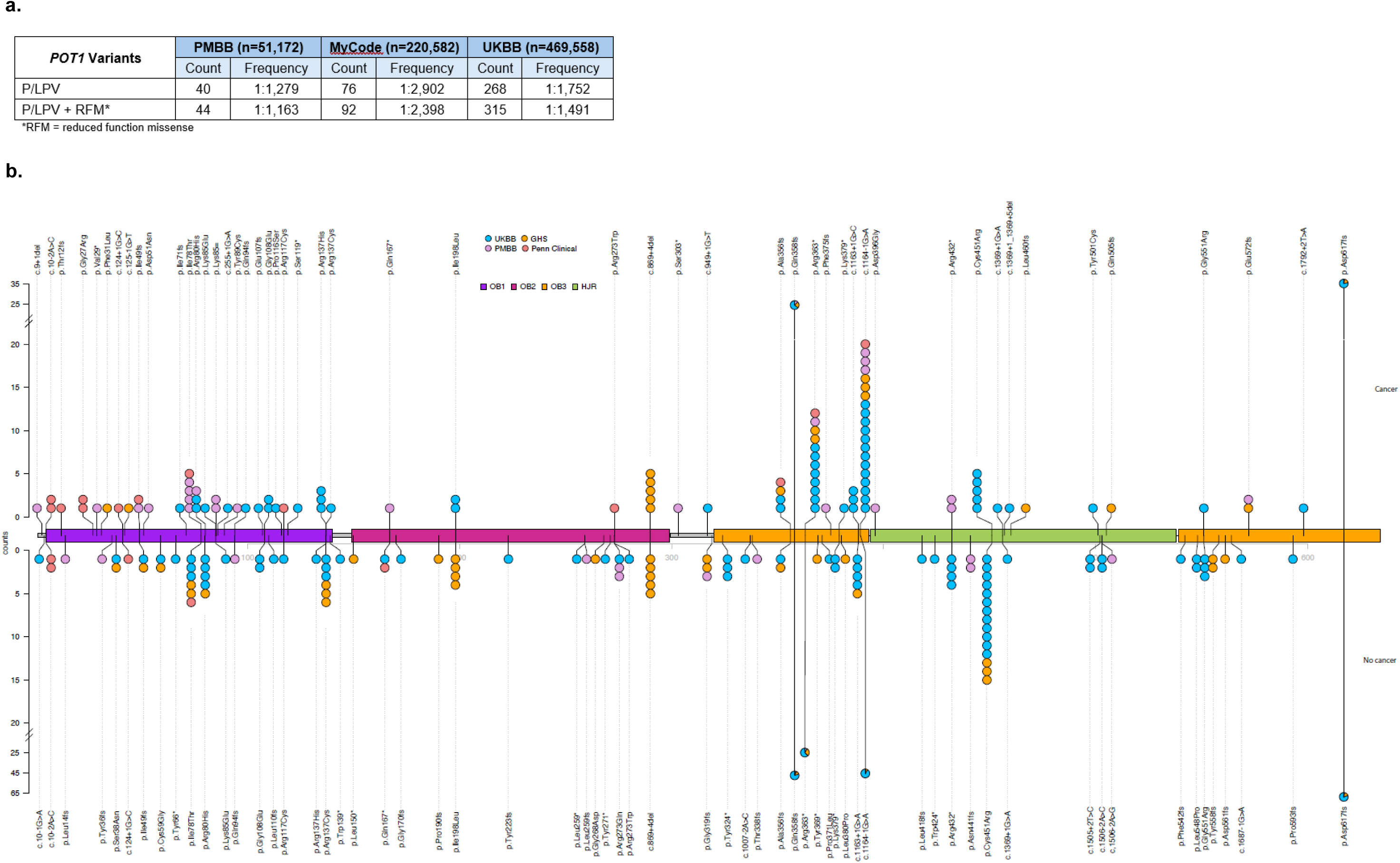
Prevalence and distribution of POT1 variants. a. Estimated prevalence of germline pathogenic/likely pathogenic *POT1* variants in the Penn Medicine Biobank (PMBB), MyCode and UK Biobank (UKBB). P/LPV = pathogenic and likely pathogenic variants, RFM = reduced function missense b. Diagram of the identified *POT1* variants annotated relative to the conserved protein domains. OB denotes Oligonucleotide/Oligosaccharide Binding (OB) fold domains. HJR denotes the embedded Holliday junction resolvase-like domain.

Nineteen of 89 variants included in this study were identified as RFM, which constituted 67/461 (14.5%) variant carriers identified across the biobank cohorts. Of the 19 missense *POT1* variants identified as RFM variants, 13/19 (68%) localize to the N-terminal DNA-binding OB1/OB2 domains, with 6/19 (32%) occurring in the C-terminal OB3/HJR region involved in TPP1 interaction and shelterin complex assembly.

### Cancer risk for individuals with POT1 variants in biobank cohorts

To further delineate the phenotype of individuals with *POT1* variants identified in an unbiased fashion, we utilized the PMBB, MyCode, and UKBB biobanks (**Supplementary Table 2**). We found a significantly increased risk of all cancers across biobanks for *POT1* P/LPVs: PMBB (OR 3.41, 95%CI 1.81-6.43, *p*=1.88x10^-4^), MyCode (OR 2.23, 95%CI 1.31-3.79, p=0.003), and UKBB (OR 1.40, 95%CI 1.07-1.84, *p*=0.015) (**Table 1**). A similar increased cancer risk was observed with the inclusion of RFM variants: PMBB (OR 3.98, 95%CI 2.15-7.37, *p*=8.20x10^-6^), MyCode (OR 1.78, 95% CI 1.08-2.95, *p*=0.024), and UKBB (OR 1.45, 95% CI 1.13-1.86, *p*=0.004) (**Supplementary Table 3**).

**Table 1:** Estimate of cancer risk for individuals with *POT1* variants (P/LPVs). Cancer risk was estimated using grouped ICD-10 codes and individual ICD-10 codes (Methods, Supplementary Methods)

| Cancer Types |  | Penn Medicine Biobank |  |  |  | MyCode |  |  |  | UK Biobank |  |  |  |
|---|---|---|---|---|---|---|---|---|---|---|---|---|---|
|  |  | Non-carriers | Carriers | Adj OR (95% CI) | Adj *p*-value | Non-carriers | Carriers | Adj OR (95% CI) | Adj *p*-value | Non-carriers | Carriers | Adj OR (95% CI) | Adj *p*-value |
|  |  | n (%) | *n* (%) |  |  | *n* (%) | *n* (%) |  |  | *n* (%) | *n* (%) |  |  |
| Total |  | 51,128 (100.0) | 40 (100.0) |  |  | 220,494 (100.0) | 74 (100.0) |  |  | 469,243 (100.0) | 268 (100.0) |  |  |
| All cancers |  | 15604 (30.5) | 24 (60.0) | 3.41 (1.81-6.43) | 1.88x10<sup>-4</sup> | 50865 (23.1) | 27 (30.7) | 2.23 (1.31-3.79) | 0.003 | 112616 (24.0) | 79 (29.5) | 1.40 (1.07-1.84) | 0.015 |
| **Grouped ICD-10 codes** | Lip, oral cavity, pharynx | 821 (1.6) | 2 (5.0) | 3.23 (0.78-13.39) | 0.135 | 1292 (0.6) | 2 (2.7) | 4.61 (1.10-19.4) | 0.036 | 2085 (0.4) | 0 (0.0) | NA | NA |
|  | Digestive organs | 2471 (4.8) | 1 (2.5) | 0.50 (0.07-3.68) | 1 | 3857 (1.7) | 1 (1.4) | 0.73 (0.10-5.34) | 0.754 | 16638 (3.5) | 9 (3.4) | 1.00 (0.51-1.95) | 0.998 |
|  | Respiratory and intrathoracic organs | 2019 (3.9) | 4 (10.0) | 2.70 (0.96-7.60) | 0.072 | 4234 (1.9) | 5 (6.8) | 3.63 (1.37-9.60) | 0.009 | 6952 (1.5) | 5 (1.9) | 1.37 (0.56-3.34) | 0.489 |
|  | Bone and articular cartilage | 387 (0.8) | 1 (2.5) | 3.36 (0.46-24.53) | 0.263 | 355 (0.2) | 2 (2.7) | 17.28 (4.19-71.30) | 8.08x10<sup>-5</sup> | 42749 (9.1) | 21 (7.8) | 0.87(0.55-1.36) | 0.544 |
|  | Melanoma and other skin | 4418 (8.6) | 6 (15.0) | 1.87 (0.78-4.45) | 0.154 | 19793 (9.0) | 9 (12.2) | 1.45 (0.68-3.09) | 0.332 | 1840 (0.4) | 2 (0.7) | 2.00 (0.50-8.05) | 0.331 |
|  | Breast | 2296 (4.5) | 1 (2.5) | 0.55 (0.07-3.97) | 1 | 7382 (3.3) | 5 (6.8) | 2.17 (0.83-5.65) | 0.112 | 19738 (4.2) | 7 (2.6) | 0.58 (0.27-1.24) | 0.158 |
|  | Female genital organs | 894 (3.5) | 1 (5.6) | 1.62 (0.22-12.21) | 0.473 | 4242 (3.2) | 2 (2.7) | 1.45 (0.35-6.07) | 0.611 | 6415 (2.6) | 3 (2.0) | 0.83 (0.26-2.60) | 0.745 |
|  | Male genital organs | 2889 (11.3) | 6 (27.3) | 2.94 (1.15-7.53) | 0.031 | 6505 (7.5) | 2 (2.7) | 0.83 (0.18-3.79) | 0.815 | 17383 (8.3) | 12 (10.2) | 1.37 (0.74-2.53) | 0.31 |
|  | Urinary tract | 2060 (4.0) | 3 (7.5) | 1.93 (0.59-6.27) | 0.218 | 3889 (1.8) | 2 (2.7) | 1.47 (0.35-6.24) | 0.598 | 7217 (1.5) | 7 (2.6) | 1.89 (0.88-4.04) | 0.101 |
|  | Eye, Brain, CNS | 537 (1.1) | 1 (2.5) | 2.42 (0.33-17.62) | 0.345 | 2042 (0.9) | 1 (1.4) | 1.44 (0.2-10.4) | 0.719 | 1679 (0.4) | 1 (0.4) | 1.06 (0.15-7.55) | 0.955 |
|  | Thyroid and other endocrine | 1382 (2.7) | 3 (7.5) | 2.92 (0.90-9.48) | 0.094 | 2746 (1.2) | 2 (2.7) | 2.17 (0.53-8.84) | 0.28 | 1201 (0.3) | 1 (0.4) | 1.43 (0.20-10.19) | 0.722 |
|  | Ill-defined, secondary, undescribed | 5826 (11.4) | 7 (17.5) | 1.65 (0.73-3.73) | 0.213 | 8784 (4.0) | 8 (10.8) | 2.98 (1.39-6.40) | 0.005 | 23972 (5.1) | 9 (3.4) | 0.66 (0.34-1.3) | 0.231 |
|  | Lymphoid and hematopoeitic | 2434 (4.8) | 4 (10.0) | 2.22 (0.79-6.25) | 0.122 | 4861 (2.2) | 5 (6.8) | 3.26 (1.29-8.23) | 0.012 | 8425 (1.8) | 24 (9.0) | 5.7 (3.73-8.72) | 9.97x10<sup>-16</sup> |
| **Individual ICD-10 codes** | Bronchus and Lung | 1705 (3.3) | 4 (10.0) | 3.22 (1.15-9.06) | 0.044 | 3744 (1.7) | 5 (6.8) | 4.30 (1.66-11.10) | 0.003 | 6217 (1.3) | 5 (1.9) | 1.5 (0.62-3.72) | 0.356 |
|  | Prostate | 2768 (10.8) | 6 (27.3) | 3.09 (1.21-7.90) | 0.026 | 6199 (7.1) | 0 (0.0) | NA | NA | 16495 (7.9) | 11 (9.3) | 1.32 (0.70-2.52) | 0.385 |
|  | CLL | 485 (0.9) | 3 (7.5) | 8.47 (2.60-27.55) | 0.007 | 1040 (0.5) | 2 (2.7) | 5.92 (1.43-24.40) | 0.014 | 1593 (0.3) | 6 (2.2) | 7.13 (3.16-16.12) | 2.30E-06 |
|  | Melanoma | 1246 (2.4) | 4 (10.0) | 4.45 (1.58-12.52) | 0.016 | 3129 (1.4) | 4 (5.4) | 4.03 (1.44-11.30 | 0.008 | 5871 (1.3) | 4 (1.5) | 1.20 (0.45-3.23) | 0.716 |
|  | Brain tumors | 405 (0.8) | 1 (2.5) | 3.21 (0.44-23.43) | 0.273 | 776 (0.4) | 0 (0.0) | NA | NA | 1227 (0.3) | 1 (0.4) | 1.44 (0.20-10.31) | 0.714 |
|  | Sarcoma | 577 (1.1) | 0 (0.0) | NA | NA | 716 (0.3) | 0 (0.0) | NA | NA | 804 (0.2) | 1 (0.4) | 2.24 (0.31-15.96) | 0.422 |
|  | Thyroid | 1301 (2.5) | 3 (7.5) | 3.11 (0.96-10.08) | 0.081 | 2073 (0.9) | 2 (2.7) | 2.86 (0.7-11.7) | 0.143 | 986 (0.2 | 0 (0.0) | NA | NA |

We evaluated association of cancer risk with *POT1* P/LPVs in both broader cancer categories and specific cancer types across biobanks (**Table 1**). In the PMBB, elevated cancer risk included the grouped ICD-10 code male genital organ cancers (OR 3.49, 95% CI 1.52-8.03, *p*=0.006); and specific codes for bronchus and lung (OR 3.22, 95% CI 1.15-9.06, *p*=0.044), prostate cancer (OR 3.09, 95% CI 1.21-7.90, *p*=0.026), CLL (OR 8.47, 95% CI 2.60-27.55, *p*=0.007) and melanoma (OR 4.45, 95% CI 1.58-12.52, *p*=0.016). In MyCode, along with an increased risk of myeloid/lymphoid malignancies (OR 3.26, 95% CI 1.29-8.23, *p*=0.012), an increased risk for lip, oral cavity, pharynx (OR 4.61, 95% CI 1.10-19.4, *p*=0.036), respiratory and intrathoracic organs (OR 3.63, 95% CI 1.37-9.60, *p*=0.009), bone and articular cartilage (OR 17.28, 95% CI 4.19-71.30, *p*=8.08x10^-5^), and unknown primary (OR 2.98, 95% CI 1.39-6.40, *p*=0.005) was observed. Increased risk for neoplasm of bronchus and lung (OR 4.30, 95% CI 1.66-11.10, *p*=0.003), CLL (OR 5.92, 95% CI 1.43-24.40, *p*=0.014), and melanoma (OR 4.03, 95% CI 1.44-11.30, *p*=0.008) was found. In the UKBB, the only grouped ICD-10 code with an elevated cancer risk was myeloid/lymphoid malignancies (OR 5.7, 95% CI 3.73-8.72, *p*=9.97x10^-16^); the specific diagnosis of CLL also was significantly increased (OR 7.13, 95% CI 3.16-16.12, *p*=2.30x10^-6^).

Including RFM variants in the analysis, the only significant associations that changed were in the PMBB dataset. Urinary tract cancers were significantly associated with *POT1* variant carrier status (OR 3.05, 95% CI 1.20-7.76, *p*=0.031), and bronchus and lung cancers were no longer significantly associated with carrier status (OR 2.90, 95% CI 1.04-8.11, *p*=0.059) (**Supplementary Table 3**).

### Detailed phenotyping of individuals with POT1 variants in Penn Medicine Biobank (PMBB) cohort

Forty-four PMBB patient-participants carried *POT1* pathogenic or likely pathogenic, or reduced function missense variants. Demographic characteristics are presented in **Supplementary Table 4**. Overall, 28/44 (63.6%) of these PMBB participants had a personal history of at least one malignancy, including 39 solid malignant neoplasms (13 types), and five hematologic malignancies (two types) (**Supplementary Tables 4, 5**). Eleven of 28 (39.3%) had multiple primary cancer diagnoses. The average number of primary solid and/or hematologic neoplasm diagnoses was 1.8 for PMBB participants with POT1 LP/P or RFM variants (**Figure 2**).

**Figure 2:**
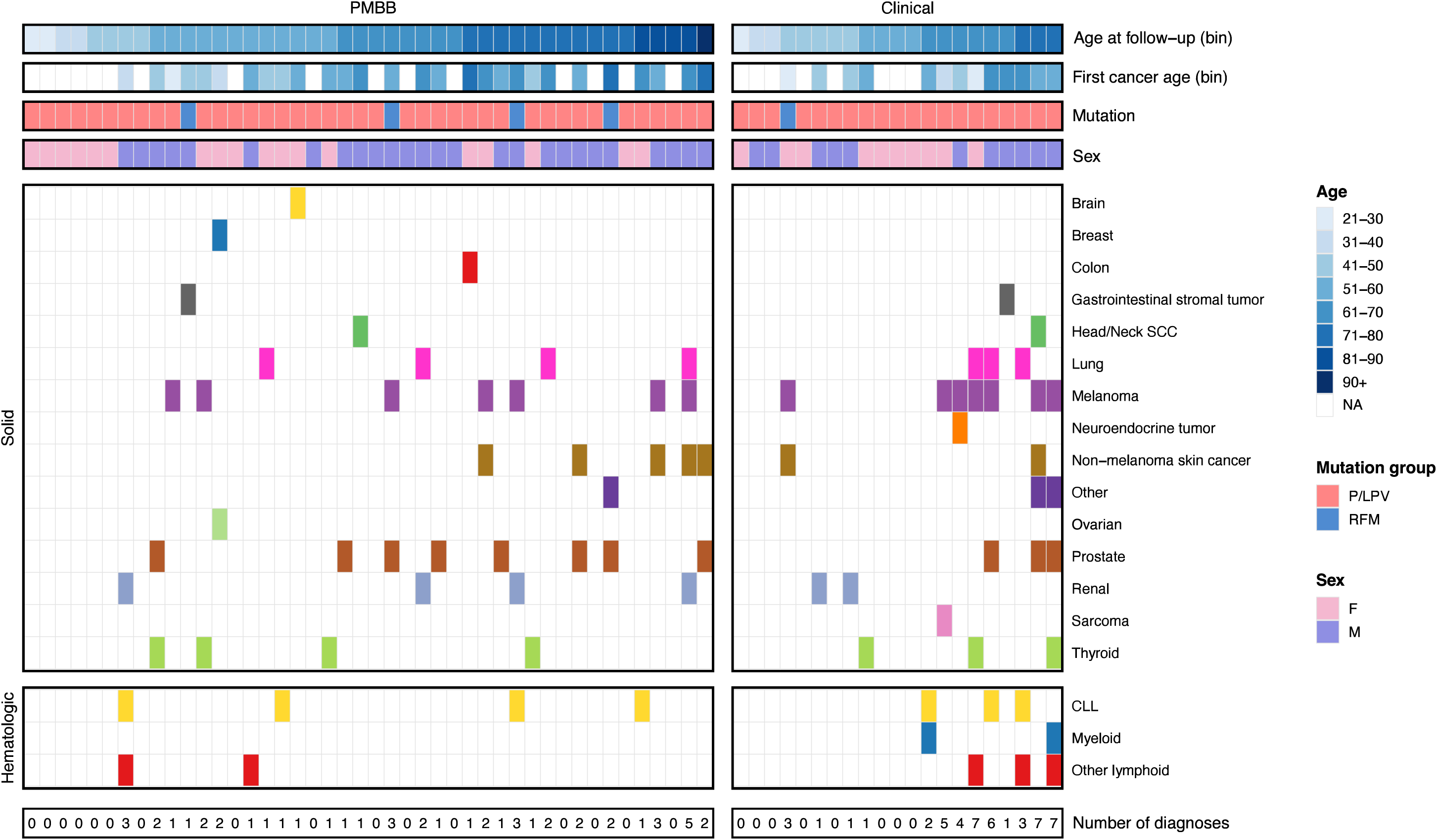
Summary of phenotypes including solid or hematologic malignancy history for individuals with *POT1* variants ascertained at Penn Medicine. P/LP = pathogenic/likely pathogenic, RFM = reduced function missense, PMBB = Penn Medicine Biobank, Clinical = individual from the clinically-ascertained Penn Medicine cohort, , M = male, F = female, SCC = squamous cell carcinoma, CLL = chronic lymphocytic leukemia.

The average age of earliest cancer diagnosis was 56.6 (SD=12.1, range 26-79) with the earliest age of cancer diagnosis melanoma between ages 26-30 years (**Figure 2**; **Supplementary Tables 5, 6**). Other cancers diagnosed before age 40 in this cohort included Hodgkin lymphoma (within age range of 26-30), CLL (within age range 36-40), renal cell carcinoma, Hodgkin lymphoma, and breast cancer (all within the age range of 36-40).

On average, unaffected individuals were younger than affected individuals at time of last assessment, 52.0 (SD=18.0) vs 65.3 (SD=11.9) years (*p*=0.003). Cancers known to be associated with *POT1* were commonly observed, including melanoma (15.9%) and CLL (9.1%). We also observed frequent diagnoses of cancers not previously described as associated with *POT1* P/LPVs, including non-small cell lung cancer (NSCLC) (9.1%; 10.8% of individuals over age 50), renal cancer (9.1%), and prostate cancer (30.8% of all males; 21.6% of males over age 50; average age diagnosis 61.9 years, SD=9.5, range 51-79) (**Supplementary Tables 5, 6**).

#### Detailed phenotyping through EHR review of adults with POT1 variants in Penn Medicine clinical cohort

Twenty-one Penn Medicine patients evaluated in a genetics clinic carried *POT1* variants with demographic characteristics presented in **Supplementary Table 4**. Thirteen individuals (61.9%) had at least one neoplasm, including 37 with solid malignant neoplasms (10 types), and eight hematologic malignancies (seven types) (**Supplementary Tables 4, 5**). Among affected individuals, 69.2% clinically ascertained had multiple primary cancer diagnoses (excluding non-melanoma skin cancer), compared to 39.3% in PMBB cohort (*p*=NS). The average number of primary solid and/or hematologic neoplasm diagnoses was 2.4 (3.7 for clinical cohort; 1.8 for PMBB participants) (**Figure 2**). The average age of earliest cancer diagnosis was 50.5 (SD=13.1, range 29-68) for the clinical cohort compared to 56.6 (SD=12.1, range 26-79) for the PMBB cohort (*p*=0.08) (**Figure 2**; **Supplementary Table 6**). The earliest age of cancer diagnosis was Hodgkin Lymphoma within age range 26-30 (**Supplementary Table 6**). The only other cancer diagnosed before age 40 in the clinical cohort was melanoma (within age range 26-30).

On average, unaffected individuals were younger than affected individuals at time of last assessment, 45.8 years (SD=11.98) vs 61.8 (SD=11.7) years (*p*=0.008). Cancers known to be associated with *POT1* P/LPs were commonly observed, including melanoma (33.3%) and CLL (14.3%). We observed the previously reported *JAK2* p.V617F variant in two of the three patients with hematological neoplasms that had somatic sequencing data available (**Supplementary Table 7**). We observed frequent diagnoses of cancers not previously described as associated with *POT1* P/LPVs, including non-small cell lung cancer (NSCLC) (14.3%; 23.1% of individuals over age 50), renal cancer (9.5%), and prostate cancer (27.3% of all males; 50% of males over age 50; one diagnosed in the age range 51-55, one in the age range of 56-60, and another in the age range of 66-70years; **Supplementary Tables 5, 6**).

In the clinical cohort, 19 individuals (90.5%) had a family history of cancer, most commonly hematologic, lung cancers and melanoma. Seven probands had additional family members who tested positive (selected pedigrees in **Supplementary Figure 2**). In families with more than one generation with individuals testing positive for *POT1* variants, consistent evidence of anticipation was not observed, as the majority did not have cases of earlier onset of cancer in successive generations.

### Cancer incidence, survival and telomere length in biobank POT1 variant carriers

Cancer incidence was calculated for individuals with *POT1* P/LPVs (**Figure 3A**) and for individuals with *POT1* P/LPVs and RFM (**Figure 3B**). In PMBB P/LPV *POT1* carriers, the cumulative risk of cancer by age 50 was 20.2% (95% CI 5.6%–32.6%) and 72.3% by age 80 (95% CI 46.5%–85.7%). With RFM included, the cumulative risk of cancer was 20.8% by age 50 (95% CI 6.8%–32.7%) and 76.7% by age 80 (95% CI 53.1%–88.4%). Carrier risk first exceeded the noncarrier estimate at age 37.0 years in both groups. For P/LPV MyCode carriers, the cumulative risk of cancer by age 50 was 13.6% (95% CI 4.3%-22.0%) and 56.9% by age 80 (95% CI 33.1%-72.3%). Similar analyses with the inclusion of RFM showed the cumulative cancer risk estimate of 11.7% by age 50 (95% CI 3.7%-19.0%) and 50.7% by age 80 (95% CI 30.8%-67.2%). Compared to the general population, cancer risk first exceeded the population-based estimate at approximately age 40 years for both groups. Lastly, in P/LPV UKBB carriers, the cumulative risk of cancer by age 50 was 3.1% (95% CI 0.9%-5.1%) and 42.3% by age 80 (95% CI 32.7%-50.5%). Compared to the general population, cancer risk first exceeded the population-based estimate at age 60.9 years. With the inclusion of RFM, the cumulative cancer risk estimate was 2.9% by age 50 (95% CI 1.0%-4.8%) and 42.9% by age 80 (95% CI 34.2%-40.4%), with cancer risk exceeding the population-based estimate at age 60.8 years.

**Figure 3:**
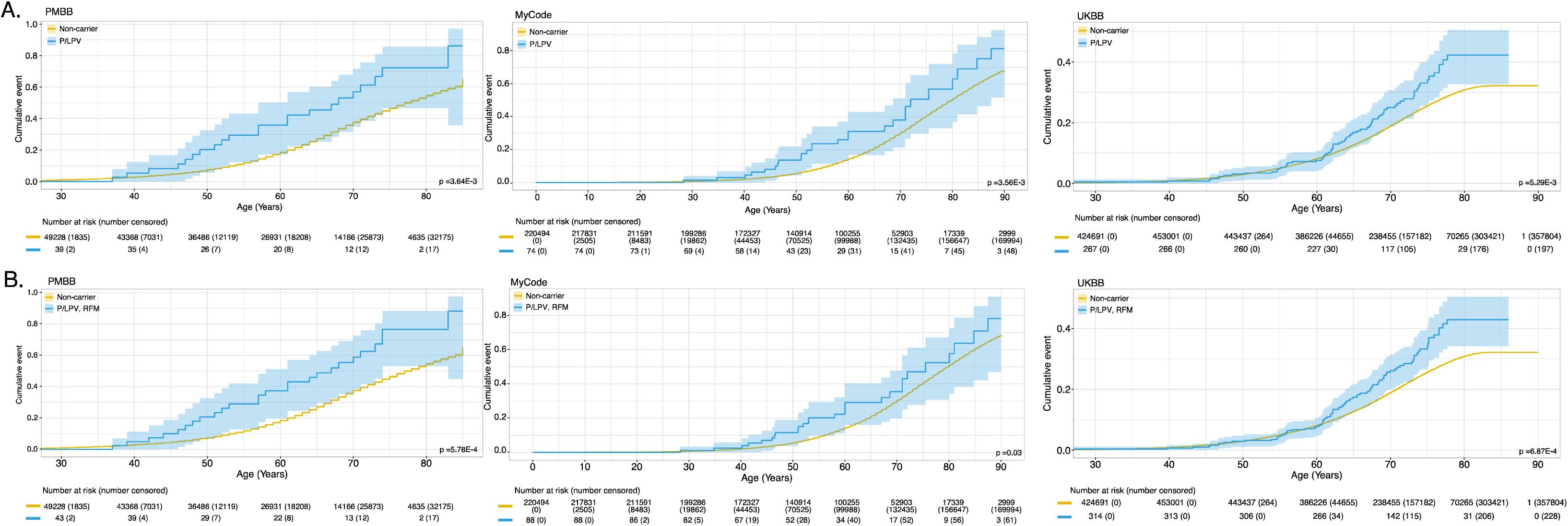
Cumulative incidence of all cancers in three biobank cohorts. a. P/LPV for PMBB (HR 3.41, 95% CI 1.81-6.43, *p*=1.88x10^-4^), MyCode (HR 2.23, 95% CI 1.31-3.79, *p*=0.003), UKBB (HR 1.40, 95% CI 1.07-1.4, *p*=0.015) b. P/LPV+ RFM for PMBB (HR 3.98, 95%CI 2.15-7.37, *p*=8.20x10^-6^), MyCode (HR 1.78, 95% CI 1.08-2.95, *p*=0.024), UKBB (HR 1.45, 95% CI 1.13-1.86, *p*=0.004).

Overall survival did not differ between *POT1* carriers and non-carriers in UKBB, including when stratified by cancer status (**Figure 4**). Additional survival analyses were run for PMBB and MyCode that also showed no difference in survival, though there were less than 20 deaths in each of these cohorts (**Supplementary Figure 3**). *POT1* variant carriers had significantly longer telomeres than non-carriers (*p*=8.25x10^-4^). We found a non-significant increase in telomere length in individuals with cancer vs. those without (p=0.66, **Figure 4**), which may be age-related as individuals with cancer were significantly older (*p*=2.25x10^-11^).

**Figure 4:**
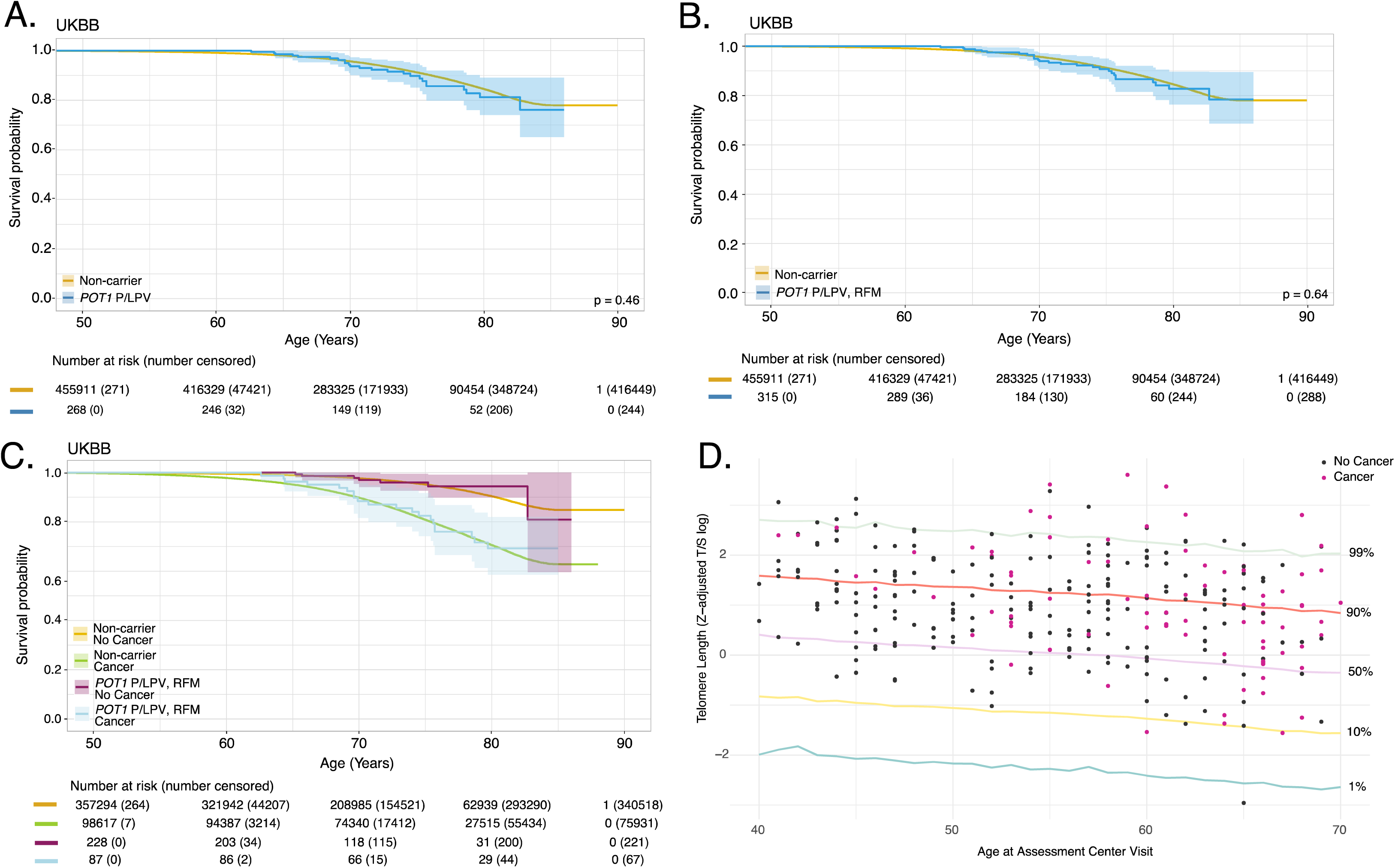
*POT1* variant carriers survival probability and telomere length in UKBB. a. P/LPV only for UKBB b. P/LPV+ RFM for UKBB. c. Survival probability of P/LPV + RFM carriers vs. non-carriers with and without cancer, UKBB d. *POT1* carriers telomere length by age at assessment and cancer status.

## DISCUSSION

We have characterized the prevalence and clinical spectrum of *POT1* germline P/LPVs in large, clinically diverse population-scale cohorts. By leveraging biobank-scale genomic data, we reduced phenotypic ascertainment bias inherent to prior clinic-based studies and provide the population-based estimates of *POT1*-associated cancer risk. In the biobank cohort evaluated here, *POT1* variants were present in approximately 1 in 1200 to 2500 individuals. The prevalence estimates suggest that *POT1* tumor predisposition may be more common than previously appreciated, and that approximately 137,000-286,000 individuals in the United States may be variant carriers. This prevalence is comparable to other autosomal dominant cancer susceptibility conditions, such as *NF1*.^38^ The higher frequency of *POT1* variants in the PMBB likely reflects both the higher proportion of Ashkenazi Jewish patients in the cohort with a known founder variant (p.Ile78Thr; 9% of Penn Medicine patients reported here),^39^ and as Penn Medicine is a tertiary referral center. Our findings emphasize the importance of considering *POT1* as part of germline genetic testing in individuals with multiple primary cancer diagnoses or a significant family history of cancer.

Across the three biobank cohorts and clinically ascertained patients, we observed a wide spectrum of cancers consistent with known *POT1* cancer associations, including melanoma, myeloid and lymphoid neoplasms, papillary thyroid cancer, and rarely glioblastoma and sarcoma. Within the biobank cohorts, *POT1-*associated tumor types that had significant associations in at least two of the cohorts studied included myeloid and lymphoid neoplasms (MyCode and UKBB), CLL (PMBB, MyCode and UKBB) and melanoma (PMBB and MyCode), confirming known associations. Although patients with other cancers suggested to be associated with *POT1* P/LP variants, including thyroid cancer, glioblastoma and sarcoma, were observed, they did not reach significance in the biobanks studied as being more common than in the non-*POT1* variant population. A previous clinical cohort study suggested that breast cancer susceptibility as part of the *POT1*-related tumor spectrum^40^; increased risk for breast cancer was not observed in our biobank analyses.

*POT1* variants were associated with an increased risk of bronchus/lung cancer in two of the three biobank cohorts; a significantly increased risk of 3.2-fold was seen in PMBB, 4.3-fold in MyCode, and non-significantly increased risk of 1.5 in the UKBB. In the PMBB cohort, we found that 9.1% of participants had bronchus/lung cancer with an average age of diagnosis of 64.2 years (SD=12.0, range 47-75) earlier the United States population average age of diagnosis of 70 years.^41^ Four of four PMBB participants with NSCLC had a history of smoking. Two prior publications also have suggested that *POT1* variants may be associated with an increased risk of lung cancer.^22,42^ We observed a variable association of *POT1* variants with prostate cancer. In the PMBB cohort, 30.8% of all males had prostate cancer. The average age of diagnosis of prostate cancer in PMBB participants was 61.9 years (SD=9.5, range 51-79) with most Gleason 6 or 7, younger than the United States average age of diagnosis of 67 years.^41^ In the UKBB, a non-significant increased risk of prostate cancer was observed, which may be due to differences in UK surveillance guidelines. However, in MyCode no men with prostate cancer were identified. A retrospective series from a commercial testing laboratory also suggested an association with prostate cancer (p=0.018, OR 2.4, 95% CI 1.14-5.06).^23^ The role of *POT1* as a susceptibility gene for lung and prostate cancers warrants further investigation in large on-going studies of these diseases as increased risk was observed in this study in some of the cohorts, although the number of cases for these cancer types was small.

Our population-based analyses support that POT1 variants are associated with moderate increase in cancer risk with odds ratios ranging from 1.40 to 3.41. A wide spectrum and high incidence of multiple cancers in individuals with *POT1* P/LPVs (69.2% of the clinical cohort) underscores the broad cancer predisposition. In PMBB and MyCode, cancer incidence curves did not diverge from non-carriers until beginning in the mid-30s; in UKBB, divergence occurred after age 60. Pediatric cancers were not reported in any individuals in the PMBB clinical or biobank cohort with deep phenotyping or observed in the MyCode cohort, suggesting that *POT1* variants are not associated with childhood cancers. These findings are similar to a case series with median age of diagnosis of first primary in the 40s, and only one of 44 individuals was reported to have a pediatric cancer.^40^ In the PMBB cohort, the earliest age of cancer diagnoses was within the age range 26-30 years (melanoma), and in the Penn Medicine clinical cohort the youngest diagnoses were at lymphoma and melanoma, both between ages 26-30 years.

The lower than anticipated overall cancer risk and lack of increased mortality rate, likely is reflective of similar age of cancer diagnosis to the general population and identification of indolent cancers, such as CLL and low-grade prostate cancer.^43,44^ A prior study suggested genetic anticipation with offspring who carried the same germline variant developing cancer several decades earlier than their parents.^45^ We did not observe clear evidence of anticipation in the seven families with multiple family members with *POT1* P/LPVs. We also did not observe a significant association between z-adjusted telomere length and cancer risk in the UKBB, recognizing that it is an imperfect surrogate for directly measuring telomere length.^46^ As telomeres reset during embryogenesis,^47^ anticipation would not be expected biologically, though telomere dynamics are still under study and may be more complex than previously appreciated. Use of telomere length to predict cancer risk in *POT1* variant carriers is not supported by our data, although further studies should be done.

We note several limitations of this study, including the reliance on structured EHR data for MyCode, requiring three instances of a diagnosis, and cancer registry data in UKBB for phenotypic information. The characteristics of each biobank cohort likely impacted frequency of *POT1* variants and disease associations. Participants in the UKBB are known to be a “healthy volunteer” population,^48^ and UKBB and MyCode cohorts have limited ancestral diversity. We did not observe some of the rare tumor types known to be associated with *POT1* variants, including cardiac angiosarcoma. Further, the number of events (deaths) in the survival analyses for PMBB and MyCode was under 20, which lead to unstable estimates of mortality. Thus, even larger sample sets of POT1 variant carriers may be needed to refine rates of associated rare cancers and mortality estimates.

Currently, providers have two vastly discrepant guidelines for the clinical management of patients with *POT1* variants; the current study provides critical data to inform these guidelines moving forward. As we found that *POT1* variants confer a moderately increased risk of cancer risk (1.5-3.5-fold) and no evidence for increased mortality, limited surveillance is a reasonable option. Given the multiple reports of *POT1* variants in melanoma cohorts,^8,9,16^ increased risk in the biobanks, and melanoma diagnoses in the 20s, our data support dermatology surveillance starting at age 18 years with consideration of surveillance every 6 months. Annual CBC starting at age 18 is also reasonable given the increased risk for hematologic malignancy observed, also as a relatively non-invasive and cost-effective test.

However, it is unknown whether early detection will change outcome.^49^ Currently, the U.S Preventative Services Task Force (USPSTF) recommends annual low dose computed tomography (LDCT) as surveillance for lung cancer in adults aged 50 to 80 with a twenty-pack year history.^50^ Our observation of NSCLC diagnoses suggest emphasizing the importance uptake of LDCT surveillance for POT1 variant carriers with a smoking history in concordance with USPSTF recommendations. Consideration also could be given to prostate specific antigen screening of men with *POT1* P/LPVs starting at age 45, consistent with guidelines for high risk men,^51, 52^ as a relatively non-invasive test, given the increased risk of prostate observed in one of the biobanks and prior studies.^17,23^ These recommendations expand upon current population-based surveillance guidelines. If more intensive surveillance is considered, several points emerge based on our data regarding timing and type. These data suggest starting surveillance at age 35 would be reasonable to consider, rather than age 18. Additionally, our data do not support a significantly increased risk of brain tumors in patients with *POT1* variants, suggesting that surveillance could be based on family history. All extensive imaging should be considered in the context of shared decision-making with patients and may depend upon their personal and family cancer history. Prospective studies to explore surveillance protocols and management guidelines are needed to optimize early cancer detection strategies and improve patient outcomes for individuals with *POT1* P/LPVs.

## Data Availability

The data that support the findings of this study are available from the corresponding author upon request, depending upon the limitations of any legal agreement.

## Author Contributions

ACR, BIS and KLN had full access to all the data in the study and take responsibility for the integrity of the data and the accuracy of the data analysis.

*Concept and design:*ACR, BIS, JK, DRS, KLN

*Acquisition, analysis, or interpretation of data:* ACR, BIS, JK, BSW, AM, AD, JH, JL, DC, RGC, PMB, DH, DRS, KLN

*Drafting of the manuscript:* ACR, BIS, KLN

*Critical review of the manuscript for important intellectual content:* all authors

*Statistical analysis:* BIS, JK, BSW, KLN

*Obtained funding:* DH, DRS, KLN

*Administrative, technical, or material support:* MR, TRH, SC, TC, AD

*Supervision:* DH, DRS, KLN

## Additional Contributions

We acknowledge the Penn Medicine BioBank (PMBB) providing data and thank the patient-participants of Penn Medicine who consented to participate in this research program. We would also like to thank the Penn Medicine BioBank team and Regeneron Genetics Center for providing genetic variant data for analysis. The authors are grateful to participants of the MyCode Community Initiative for use of their genomic clinical data, without whom parts of the study are not feasible. We thank the Geisinger-Regeneron Genetics DiscovEHR Collaboration for access to the genotype and phenotype data. We also acknowledge that this work used the computational resources of the NIH High Performance Computing Biowulf cluster. We thank Jenny Li for assistance with data visualization.

## Nonauthor Collaborators (Penn Medicine Biobank)

Daniel J. Rader, Marylyn D. Ritchie, JoEllen Weaver, Nawar Naseer, Giorgio Sirugo, Afiya Poindexter, Jenna Dever, Aidan Harvey, Sydney Linn, Naman Srivastava, Meghan Livingstone, Fred Vadivieso, Stephanie DerOhannessian, Teo Tran, Julia Stephanowski, Salma Santos, Ned Haubein, Joseph Dunn, Anurag Verma, Colleen Morse Kripke, Marjorie Risman, Renae Judy, Colin Wollack, Shefali Verma, Scott Damrauer, Yuki Bradford, Scott Dudek, Theodore Drivas, Zachary Rodriguez.

## Ethics Declaration

This study was approved by the University of Pennsylvania IRB, protocol #813913. This research has been conducted using the UK Biobank Resource under Application Number 54389. Geisinger data was collected and analyzed under the Geisinger IRB approved protocol #2021-0818.

## Disclaimer

This research was supported in part by the Intramural Research Program of the National Institutes of Health (NIH). The contributions of the NIH authors are considered Works of the United States Government. The findings and conclusions presented in this paper are those of the author(s) and do not necessarily reflect the views of the NIH or the U.S. Department of Health and Human Services.

