## Supplementary online materials for "Cancer risk and survival outcomes in germline *POT1* variant carriers"

**Supplementary Methods**	Pages 2-4

**Supplementary Figure 1.** Allele depletion studies for p.Asp617fs	Page 5

**Supplementary Figure 2:** Representative, partial pedigrees	Page 6

**Supplementary Figure 3**. Survival analyses for PMBB and Geisinger MyCode	Page 7

**Tables (Excel)**

**Supplementary Table 1 (ST1).** *POT1* variants included in the current study

**Supplementary Table 2 (ST2).** Demographics of the Biobanks - UK Biobank, Geisinger Health System, Penn Medicine Biobank

**Supplementary Table 3 (ST3).** Cancer Associations – POT1 P/LPV + RFM

**Supplementary Table 4 (ST4).** Demographics of the Penn Medicine Biobank and Clinical Cohorts

**Supplementary Table 5 (ST5).** Overview of Cancer Diagnoses in Penn Medicine POT1 variant carriers

**Supplementary Table 6 (ST6).** Malignant and Benign Neoplasms for Each Penn Medicine Patient

**Supplementary Table 7 (ST7).** Results of Somatic Genetic Testing in Hematological Neoplasms

**Supplementary Method****s**

Allele depletion studies

log2 fold-change in allele frequency over three weeks of screening was calculated for each individual allele present in three biological replicates. Significant depletion and enrichment were determined by wilcoxon rank sum comparison to the 180 synonymous or frame-shift alleles closest to the same week one allele frequency, respectively. Alleles that were not significantly depleted compared with the synonymous mutations were classified as persistent. Alleles that were not significantly enriched compared with the frameshift alleles were classified as depleted. Alleles significantly different from both were classified as hypomorphs^30^.

Demographic and clinical information data collection

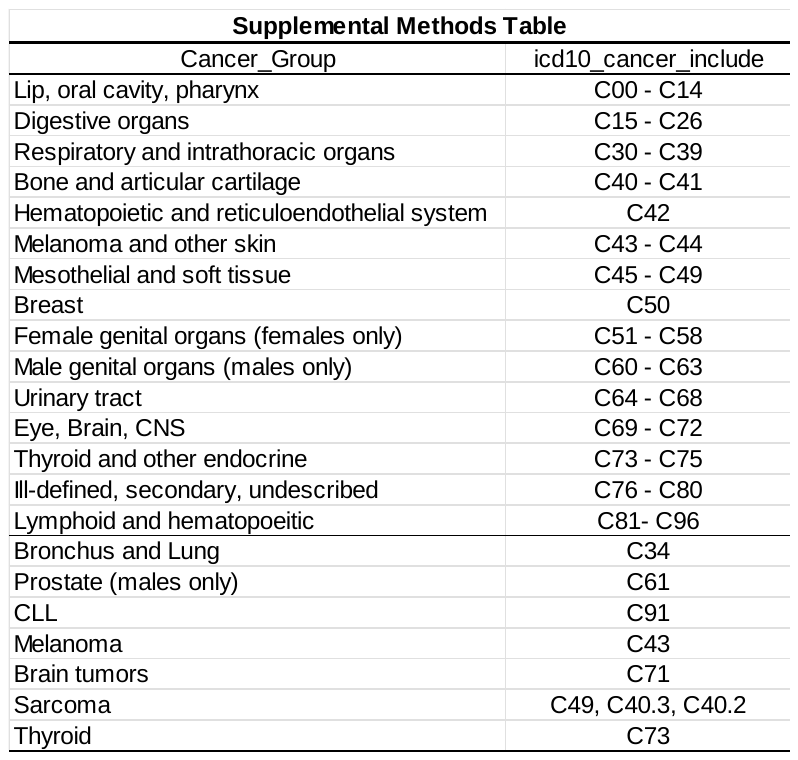
The following demographic and clinical information were collected: sex, age, race, ethnicity, smoking history, genetic test results, past medical history, clinical cancer diagnosis, age at cancer diagnosis, cancer stage, cancer pathology, family history. Data were collected from clinic notes, pathology reports, and pedigrees. For PMBB, ICD-9 and ICD-10 codes were used, with at least two instances of each code needed for diagnosis. For UKBB, fields 41270/41271 (Diagnoses – ICD10/9), 40001 (Underlying cause of death), and 40006/40013 (Type of cancer: ICD10/9) were extracted on 7/12/2024. For MyCode, the EHR and the Geisinger Cancer Registry were queried. For the cancer association studies, tumors were done based on the ICD10 codes shown in Supplemental Methods Table. Individuals needed to have two instances of each code to be included.

Bioinformatics analysis: Data preprocessing

For the VCFs (UKBB field 23157) sample level filtering for GQ<30, ABHet<0.3, DP <5 was done.

Bioinformatics analysis: Calculation of telomere length from UKBB

Telomere lengths were extracted from UKBB fields 22192 (Z-adjusted T/S log) and age at recruitment were extracted from field 21003. Scatter plot was plotted using ggplot2.

Statistical Analysis

Continuous variables are described as mean (standard deviation) and range; categorical variables are expressed as frequency and percentage. Two-tailed Fisher’s exact tests and Mann-Whitney U tests were performed to test statistical differences in demographics and frequency of multiple cancer diagnoses between clinical and biobank cohorts (at 𝛼=0.05 level). Case control analyses were performed using R version 4.1.0 and SAS Enterprise Guide v8.3.0.013. Demographic comparison was performed using t-test for age, BMI, Fisher’s exact test for sex, smoking history, cancer, and death. Cancer risk odds ratio was modeled using logistic regression with year of birth, sex, smoking history and BMI as covariates. Overall survival and time-to-cancer were plotted using the survival and ggplot2 packages and cox-proportional hazard model was modeled with year of birth, sex, smoking history, genetically derived ancestry and BMI as covariates. Survival analysis could not be performed for the MyCode or PMBB cohorts due to events (deaths) less than 20. Lollipop plot was generated in R using ggplot2.

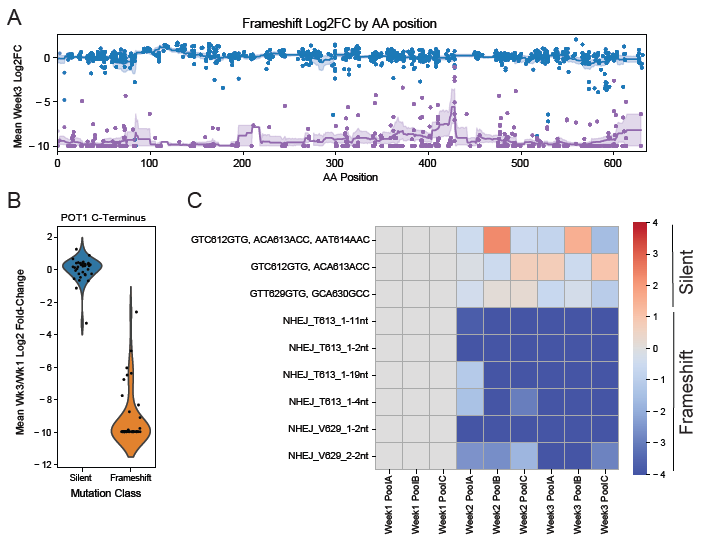

**Supplementary Figure 1: Allele depletion studies for p.Asp617fs.** (A) Analysis of DMS data generated in POT1 loHAP cells (Martin et al, PMID:40015989). Shown are log2-fold changes in allele frequency between week 3 and week 1 of the screen for frameshift mutations (purple) compared to synonymous mutations (blue) across the POT1 coding sequence. (B) Violin plot summarizing allele-frequency changes for synonymous (blue) and frameshift (orange) mutations located within the last coding exon of POT1 as shown in (A). (C) Heatmap depicting allele-frequency changes for synonymous and frameshift mutations before and after amino acid 617 in POT1 across three biological replicate allele pools

**Family 1**	**Family 2**

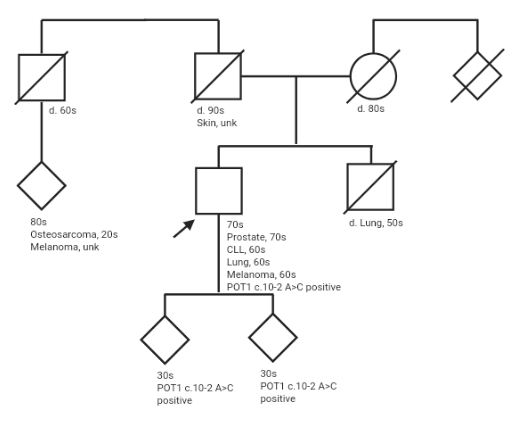

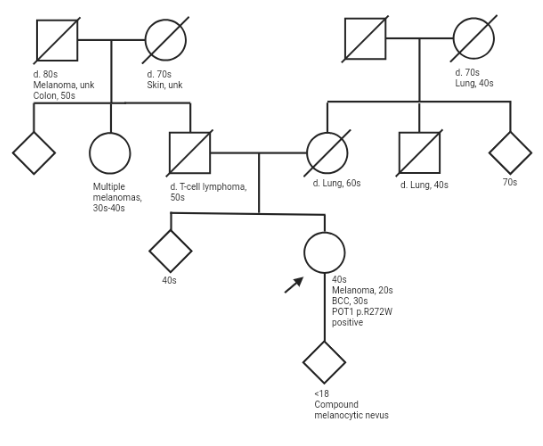

**Family 3**	**Family 4**

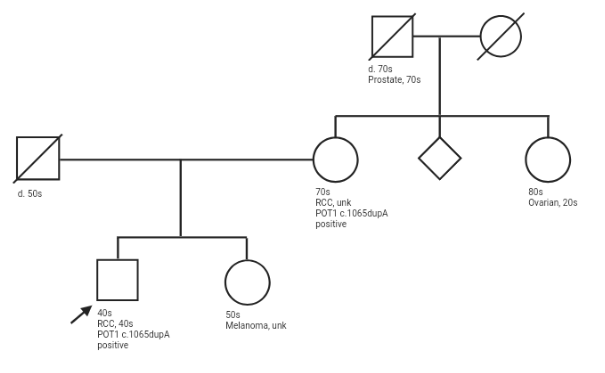

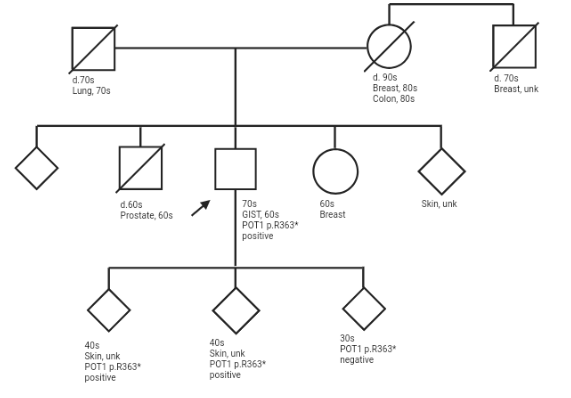

**Supplementary Figure 2: Representative, partial pedigrees.** Not all family members are included in these pedigrees, and sex has been omitted for some family members.

**Supplementary Figure 3: *POT1* variant carriers survival probability in PMBB and MyCode**

**A.**

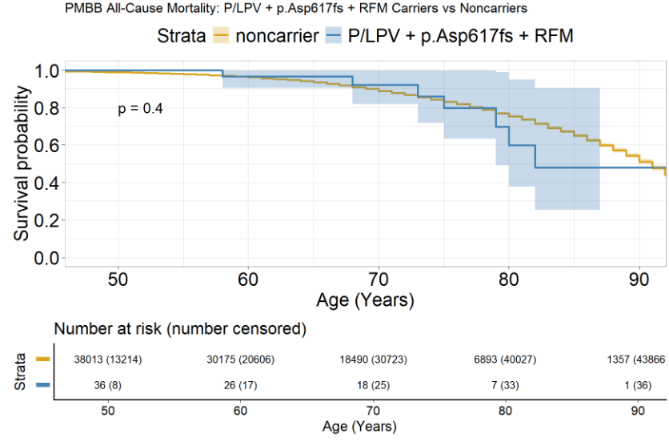

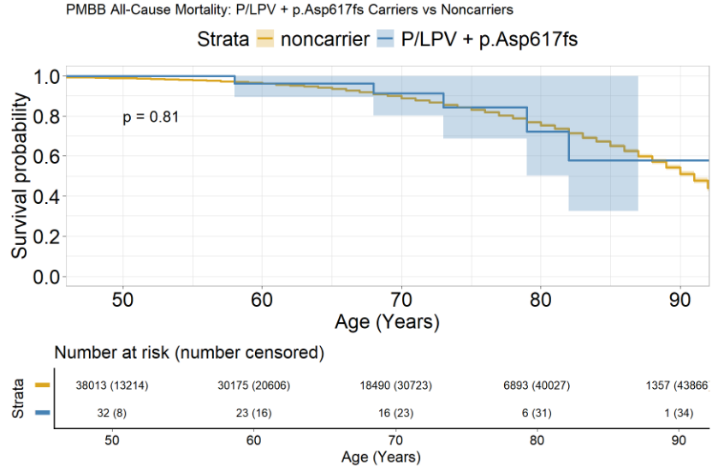

**B. MyCode**

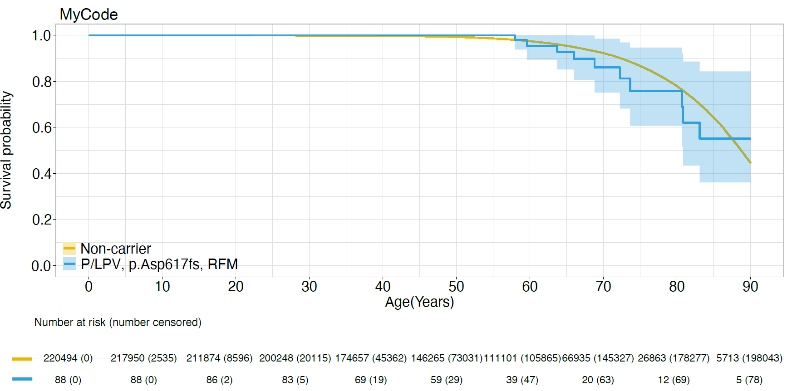

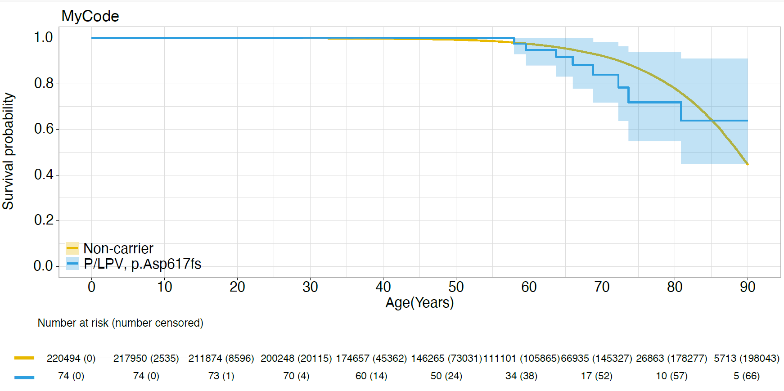
